# Multiday cycles of heart rate are associated with seizure likelihood

**DOI:** 10.1101/2020.11.24.20237990

**Authors:** Philippa J. Karoly, Rachel E. Stirling, Dean R. Freestone, Ewan S. Nurse, Matias Maturana, Amy Halliday, Andrew Neal, Nicholas M. Gregg, Benjamin Brinkmann, Mark P. Richardson, Sonya B. Dumanis, Andre La Gerche, David B. Grayden, Wendyl D’Souza, Mark J. Cook

**Affiliations:** Department of Biomedical Engineering, The University of Melbourne; Graeme Clark Institute for Biomedical Engineering, The University of Melbourne; Departments of Medicine and Neurology, The University of Melbourne, St Vincent’s Hospital, Melbourne; Seer Medical, Australia; Bioelectronics Neurophysiology and Engineering Lab, Department of Neurology, Mayo Clinic, Rochester, MN; Division of Neuroscience, King’s College London, London, UK; Epilepsy Foundation, Landover, Maryland; Sports Cardiology Laboratory, Baker Heart & Diabetes Institute, Melbourne, Australia

## Abstract

Circadian and multiday rhythms are found across many biological systems, including cardiology, endocrinology, neurology, and immunology. In people with epilepsy, epileptic brain activity and seizure occurrence have been found to follow circadian, weekly, and monthly rhythms. Understanding the relationship between these cycles of brain excitability and other physiological systems can provide new insight into the causes of multiday cycles. The brain-heart link is relevant for epilepsy, with implications for seizure forecasting, therapy, and mortality (i.e., sudden unexpected death in epilepsy).

We report the results from a non-interventional, observational cohort study, Tracking Seizure Cycles. This study sought to examine multiday cycles of heart rate and seizures in adults with diagnosed uncontrolled epilepsy (N=31) and healthy adult controls (N=15) using wearable smartwatches and mobile seizure diaries over at least four months (M=12.0, SD=5.9; control M=10.6, SD=6.4). Cycles in heart rate were detected using a continuous wavelet transform. Relationships between heart rate cycles and seizure occurrence were measured from the distributions of seizure likelihood with respect to underlying cycle phase.

Heart rate cycles were found in all 46 participants (people with epilepsy and healthy controls), with circadian (N=46), about-weekly (N=25) and about-monthly (N=13) rhythms being the most prevalent. Of the participants with epilepsy, 19 people had at least 20 reported seizures, and 10 of these had seizures significantly phase locked to their multiday heart rate cycles.

Heart rate cycles showed similarities to multiday epileptic rhythms and may be comodulated with seizure likelihood. The relationship between heart rate and seizures is relevant for epilepsy therapy, including seizure forecasting, and may also have implications for cardiovascular disease. More broadly, understanding the link between multiday cycles in the heart and brain can shed new light on endogenous physiological rhythms in humans.

## Introduction

Cyclic phenomena are ubiquitous in biological systems. In the field of chronobiology, circadian rhythms (and related 24-hour rhythms) have been widely studied. However, other timescales, including weekly (circaseptan), monthly (circalunar or circatringian), seasonal and even longer rhythms have also been observed across a diverse range of physiological functions.^1–3^ In neurology, huge strides have been made in chronic brain recording devices, leading to overwhelming evidence that multiday cycles govern brain excitability in people with epilepsy.^4–7^

The phenomena of multiday cycles in epilepsy were identified through observation of individuals’ seizure patterns.^7^ Subsequent studies using chronic electroencephalography (EEG) have shown that periodicity of seizure occurrence is underpinned by individual-specific circadian and multiday rhythms of epileptic activity^5,8^ and brain excitability^6^ in humans and other mammals.^9,10^ Importantly, multiday cycles in epilepsy exist for most people,^4–6,11^ and appear to be ‘free-running’ in the sense that they are not tied to environmental cues (weekday, lunar cycle, calendar),^12^ are equally prevalent in men and women^4,5,7,11^ and are observed across epilepsy syndromes and seizure types.^4^

Although multiday cycles of brain activity have been predominantly investigated in people with epilepsy, it is unlikely that these slower rhythms are limited to epilepsy. Other episodic psychiatric conditions are suggestive of multiday modulation, including bipolar disorder,^13^ depression^14^ and other psychopathologies.^15^ Aside from neurology, multiday cycles are recognised in cardiology, immunology and endocrinology, for instance.^3^ Therefore, we hypothesise that free-running multiday rhythms are widespread across major organ systems, analogous to circadian rhythms. However, identifying systemic, multiday oscillatory biomarkers has been limited by availability of chronic recording capabilities.

Of the wider physiological systems, long-term cardiac monitoring is more accessible than other physiological monitoring pertinent to multiday cycles. Indeed, chronobiology has a long history in cardiology and the aetiology of heart disease. In addition to well documented circadian rhythms of cardiac electrophysiology and arrhythmias,^16^ some studies have also identified intrinsic multiday cycles of cardiac output.^17–19^ These studies suggest that longer rhythms also drive cardiac activity, which may be linked to aforementioned multiday rhythms in the brain.

Brain-heart dynamics have long been of interest in epilepsy research and clinical management. Epileptic seizures can cause functional changes in the autonomic nervous system, often detected as acute changes in heart rate (tachycardia, bradycardia) or heart rate variability (HRV) near the onset or offset of seizures.^20,21^ Pre-ictal changes in heart rate and HRV have been identified as potential triggers or predictors of impending seizures, ^21^ however to our knowledge, the relationship between heart rate changes and seizures has not been investigated over multiday timescales.

Understanding the relationship between the brain and heart at slower timescales (e.g., weekly, monthly or seasonal) can provide new insight into causes of multiday physiological cycles. If cardiac electrophysiology shows multiday rhythms akin to cycles of brain excitability, this will provide a new avenue to monitor aspects of neurological diseases. Similarly, characterising slow changes in cardiac activity may be important for treating heart conditions. This study aimed to 1) identify possible multiday cycles of heart rate, and 2) determine whether there are associations between heart rate cycles and seizure likelihood in people with epilepsy.

## Results

Descriptive statistics of continuous data are reported as mean +/- standard deviation. We defined ultradian cycles as rhythms less than 24 hours, circadian cycles as 24-hour rhythms and multiday cycles as any rhythm greater than 24 hours. We also defined the following subsets of multiday cycles: ‘about-weekly’ as 5 – 9 days and ‘about-monthly’ as 28 – 32 days.

### Participants

There were 31 participants with epilepsy (21 female) with 12.0 +/- 5.9 months of continuous heart rate data at 89.3 +/- 8.2% adherence. There were 15 healthy control participants (6 female) with 10.6 +/- 6.4 months of continuous heart rate recorded at 89 +/- 7% adherence. For patients with epilepsy, the cumulative seizure diary duration was 55.0 years (mean 21.3 +/- 21.0 months), documenting over 3,619 seizures (mean 117 +/- 118 seizures) with 2,244 of these (mean 72 +/- 100) reported during the wearable monitoring period.

Table 1 shows the demographics and statistics of participants with epilepsy. Participants’ anti-epileptic drugs (AED) are given in Supplementary Table 1. Healthy control participants are shown in Supplementary Table 2.

**Table 1.** Demographics of participants with epilepsy.

| Participant | Gender | Epilepsy Type<sup>a</sup> | Recording length (months) | Seizures | Adherence (%) | Average Heart Rate (BPM<sup>b</sup>) |
| --- | --- | --- | --- | --- | --- | --- |
| P1 | F | Focal (T) | 16.5 | 105 | 100 | 93 |
| P2 | F | GGE | 12.7 | 3 | 97 | 70 |
| P3 | M | Focal (T) | 14.5 | 213 | 88 | 79 |
| P4 | M | GGE | 5.6 | 65 | 92 | 80 |
| P5 | M | Focal (TO) | 14.8 | 120 | 94 | 65 |
| P6 | F | Multi-focal | 4.0 | 37 | 80 | 89 |
| P7 | F | Focal | 20.6 | 29 | 100 | 78 |
| P8 | F | GGE (JME) | 4.1 | 2 | 80 | 70 |
| P9 | F | Focal (HH) | 13.2 | 0 | 99 | 62 |
| P10 | M | Focal | 13.4 | 67 | 100 | 70 |
| P11 | F | Focal (T) | 19.3 | 32 | 99 | 69 |
| P12 | F | Focal (HH) | 5.2 | 0 | 80 | 77 |
| P13 | F | GGE (JAE) | 6.9 | 169 | 80 | 82 |
| P14 | M | Focal (T) | 20.2 | 24 | 82 | 75 |
| P15 | M | Focal | 4.8 | 20 | 80 | 84 |
| P16 | M | Focal (TP) | 6.9 | 29 | 80 | 76 |
| P17 | F | Focal (T) | 6.2 | 7 | 80 | 69 |
| P18 | F | Multi-focal | 10.8 | 5 | 80 | 74 |
| P19 | F | Focal (T) | 21.8 | 103 | 82 | 87 |
| P20 | F | Focal (T) | 21.3 | 7 | 94 | 77 |
| P21 | M | Focal (T) | 8.1 | 21 | 93 | 72 |
| P22 | F | Focal (T) | 9.3 | 0 | 99 | 85 |
| P23 | F | Focal (T) | 22.3 | 416 | 99 | 73 |
| P24 | F | Multi-focal | 14.7 | 4 | 92 | 62 |
| P25 | M | GGE (JME) | 6.2 | 3 | 80 | 64 |
| P26 | F | GGE (JAE) | 4.2 | 14 | 98 | 70 |
| P27 | F | Focal (T) | 13.6 | 148 | 80 | 86 |
| P28 | F | Focal | 10.8 | 6 | 97 | 79 |
| P29 | F | Focal (F) | 9.0 | 251 | 89 | 78 |
| P30 | F | Multi-focal | 15.8 | 58 | 83 | 88 |
| P31 | M | Focal and Generalized (DEE) | 16.5 | 286 | 92 | 62 |
|  | **21 Female**<br>**10 Male** |  | **M=12.0**<br>**SD=5.9** | **M=72.4**<br>**SD=100.8** | **M=89.3**<br>**SD=8.2** | **M=75.6**<br>**SD=8.5** |

<sup>a</sup>Epilepsy types are separated into Focal, Multi-focal, GGE or Focal and Generalised. The epilepsy syndrome or lobar epileptogenic zone is presented in brackets where the data is available. T = temporal, TO = temporo-occipital, TP = temporo-parietal, HH = hypothalamic hamartoma, DEE = Developmental and Epileptic Encephalopathy, JME=juvenile myoclonic epilepsy, JAE=juvenile absence epilepsy, GGE=genetic generalised epilepsy).
<sup>b</sup>Beats per minute

### Heart rate cycles in people with epilepsy

We first investigated whether multiday cycles of average heart rate were present in people with epilepsy. At least one heart rate cycle was found in all participants in the cohort. Cycles were found at weekly, monthly, and longer timescales (see Figure 1b). 94% (N=29/31) had a multiday cycle, 55% (N=17/31) demonstrated about-weekly cycles and 29% (N=9/31) demonstrated about-monthly cycles (Table 2). Only 9 of the 17 about-weekly cycles were found at precisely 7 days, suggesting that about-weekly cycles were generally not driven by the artificial working week (Supplementary Figure 7). 23% of participants (N=7/31) also had a shorter ultradian cycle. The distributions of cycles were similar for men (Figure 1c) and women (Figure 1d), including about-monthly cycles (p = 0.62 using Kolmogorov-Smirnov test for equivalence). Only two participants’ about-monthly cycles were found at precisely 29.5 days (the lunar period).

**Table 2.**
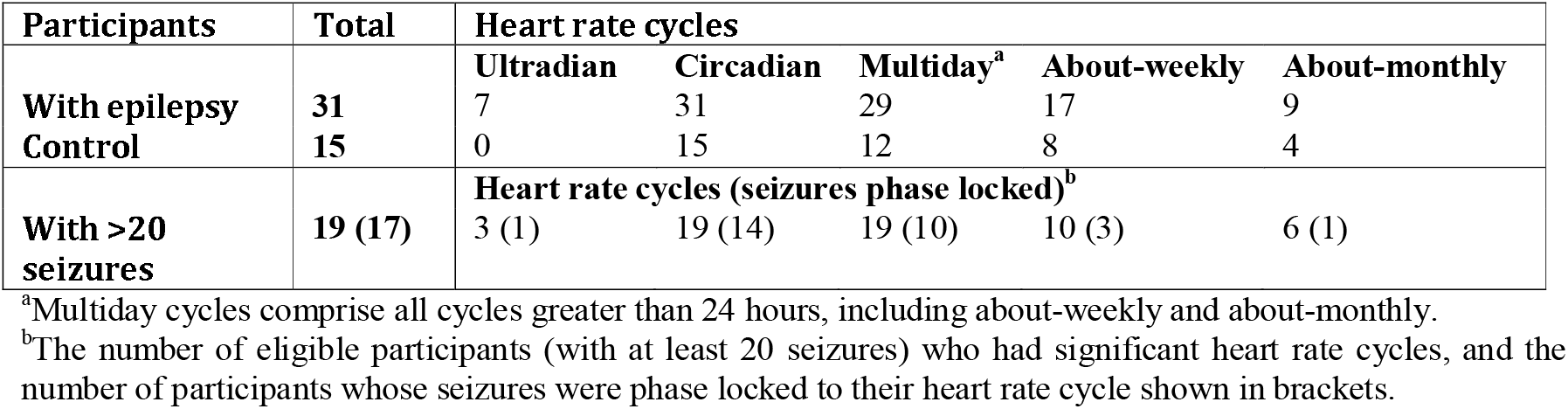
Participants with significant heart rate cycles and phase locking of seizures.

**Figure 1.**
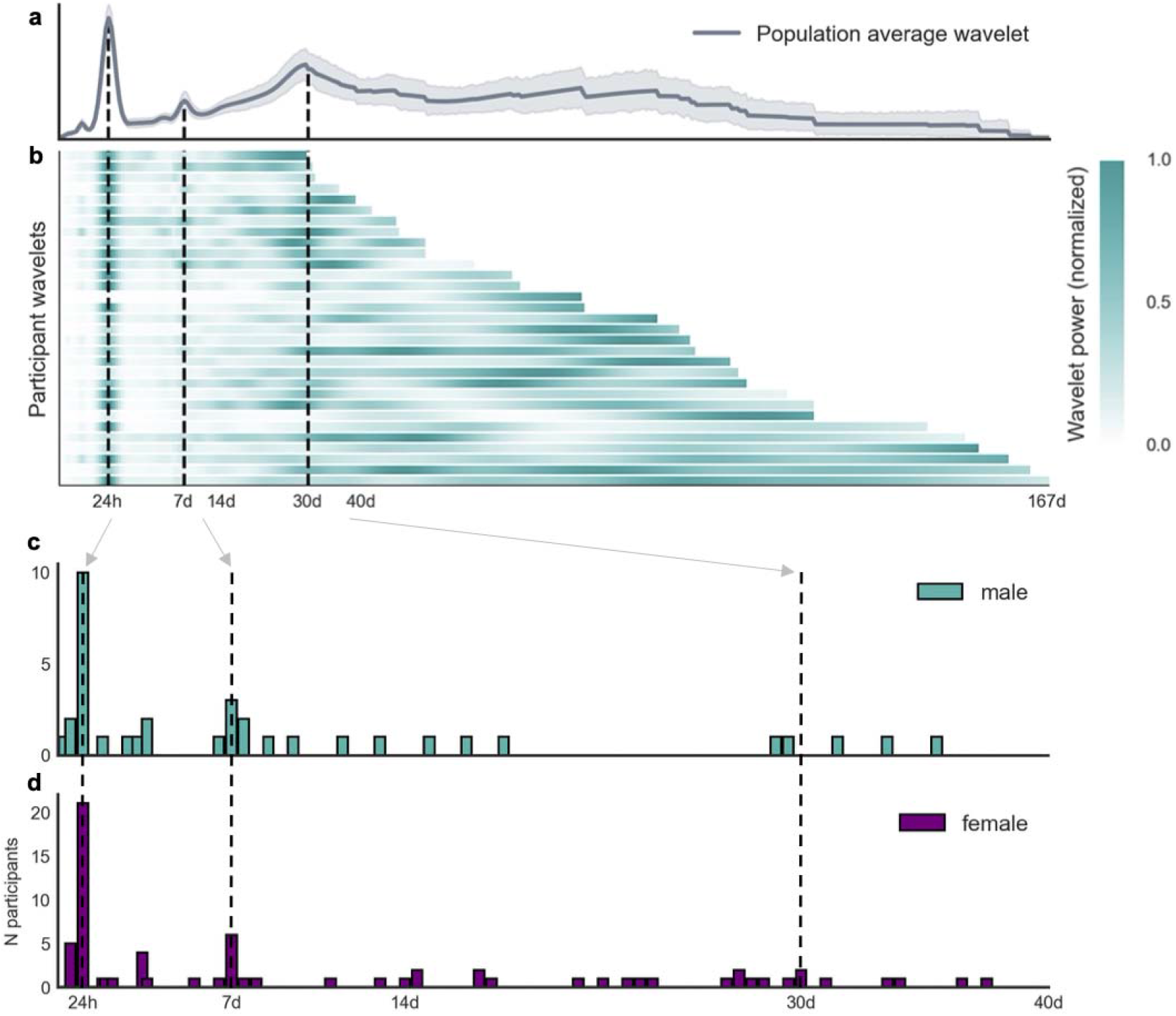
Distribution of heart rate cycles. **(A)** Cycle strength (expressed as the normalised wavelet power, y-axis) for different periods (x-axis, logarithmic scale) averaged across the cohort. Note that wavelet power was normalized between 0 and 1 (by subtracting the minimum and dividing by the range) for each participant to facilitate visualization. (**B)** Raster plot showing cycle strength (colour bar) for each individual (y-axis) at different periods (logarithmic scale). (**C, D)** Number of people (y-axis) with significant cycles at different periods up to 40 days (x-axis) for men and women, respectively. Note that the x-axis (up to 40 days) is a subset of the x-axis in panels A and B (up to 167 days) as indicated by the grey arrows and black dotted lines.

When averaged across the entire cohort, a clear peak at 24 hours was observed, as well as smaller peaks at around one week and one month (Figure 1a) (see Supplementary Figures 3,4 for individual analyses).

Striking examples of multiday heart rate cycles for two participants are shown in Figure 2. Cycles are apparent from visual inspection of average heart rate (Figure 2a,d) and were robust over the duration of recording (Figure 2b,e). Wavelet analysis confirmed significant cycles at daily (24 h), about-weekly (7 d) and multiday (15.0 and 33.5 d) periods for P21 (Figure 2c), and daily (24 h) and about-monthly (30.5 d) periods for P30 (Figure 2f).

**Figure 2.**
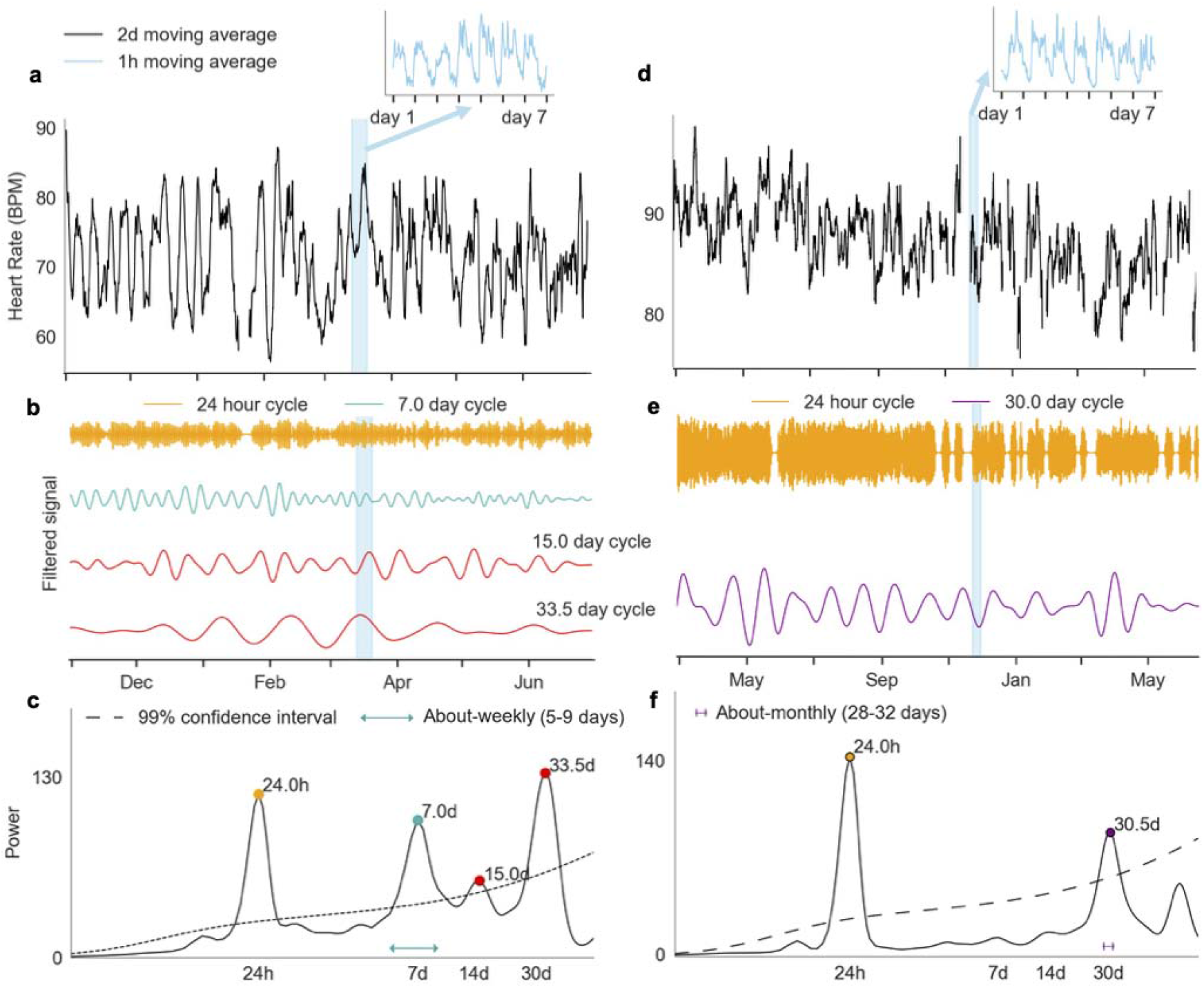
Examples of multiday heart rate cycles. Data are shown for two different participants, P21 (a-c) and P30 (d-f). **(A, D):** Heart rate (y-axis) smoothed with a 2-day moving average filter shows multiday cycles. Insets (blue) show circadian rhythms of heart rate. **(B, E):** A graphical representation of the bandpass filtered heart rate signals for different cycles (corresponding respectively to spectrum peaks in panels **(C, F)**. Note that the signal amplitudes for different cycles (coloured traces) have been normalised to the same range. **(D, G):** Wavelet power spectra for different scales (x-axis). Significant cycle periods (peaks) are labelled with coloured dots.

### Heart rate cycles in participants without epilepsy

Heart rate cycles were investigated in healthy controls. Figure 3 shows the distribution of multiday heart rate cycles across the control cohort (two individual examples are shown in Supplementary Figure 5). Although it is difficult to draw conclusions from a small cohort, cycles did appear to be more common at daily and weekly timescales, evident from the peaks in the population average (Figure 3a). Out of the total cohort of 15 people (6 female), all had a circadian cycle, 12 had a multiday cycle, with 8 showing about-weekly cycles and 4 showing about-monthly cycles (Table 2). All four of the about-monthly cycles occurred in women; although, the overall distributions of cycles were not significantly different between men and women (p = 0.51 using Kolmogorov-Smirnov test for equivalence).

**Figure 3.**
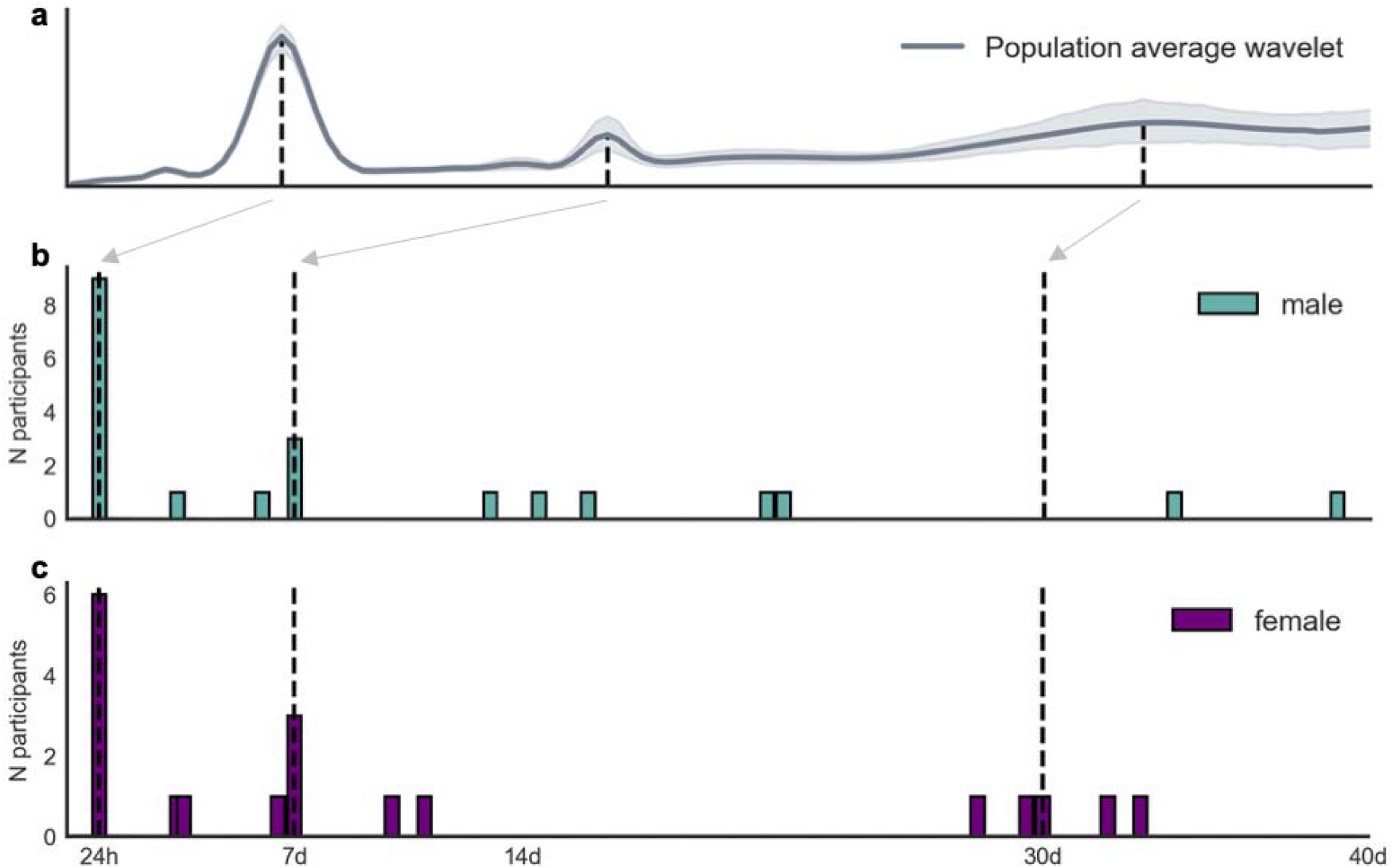
Distribution of heart rate cycles in people without epilepsy. **(A):** Cycle strength (expressed as the normalised wavelet power, y-axis) for different periods (x-axis, logarithmic scale) averaged across the cohort. (**B, C):** Number of people (y-axis) with significant cycles at different periods up to 40 days (x-axis) for men and women, respectively. The grey arrow and black dotted lines show 24-hour, 7-day and 30-day locations along the x-axes.

### Multiday heart rate cycles related to seizure risk

Among the 31 participants with epilepsy, 19 (10 female) had recorded at least 20 seizures during the wearable recording time and, therefore, were eligible for further seizure analysis. Phase locking of seizures to heart rate cycles was quantified by the synchronisation index (SI, see Methods). Of these eligible participants, 17 (89%) had seizures significantly locked onto at least one heart rate cycle (see Table 2). Eight people had both fast (circadian or ultradian) and multiday cycles comodulated with seizure likelihood, 10 had seizures significantly locked onto a multiday cycle and 14 had seizures significantly locked onto a circadian cycle.

Figure 4 shows three different example participants whose seizures were significantly synchronised to their underlying multiday heart rate cycles, demonstrated by the tight distributions of seizures (SI values between 0.44 – 0.56) with respect to these individuals’ circadian (Figure 4b), about-weekly (Figure 4d) and 14.5-day multiday (Figure 4f) cycles.

**Figure 4.**
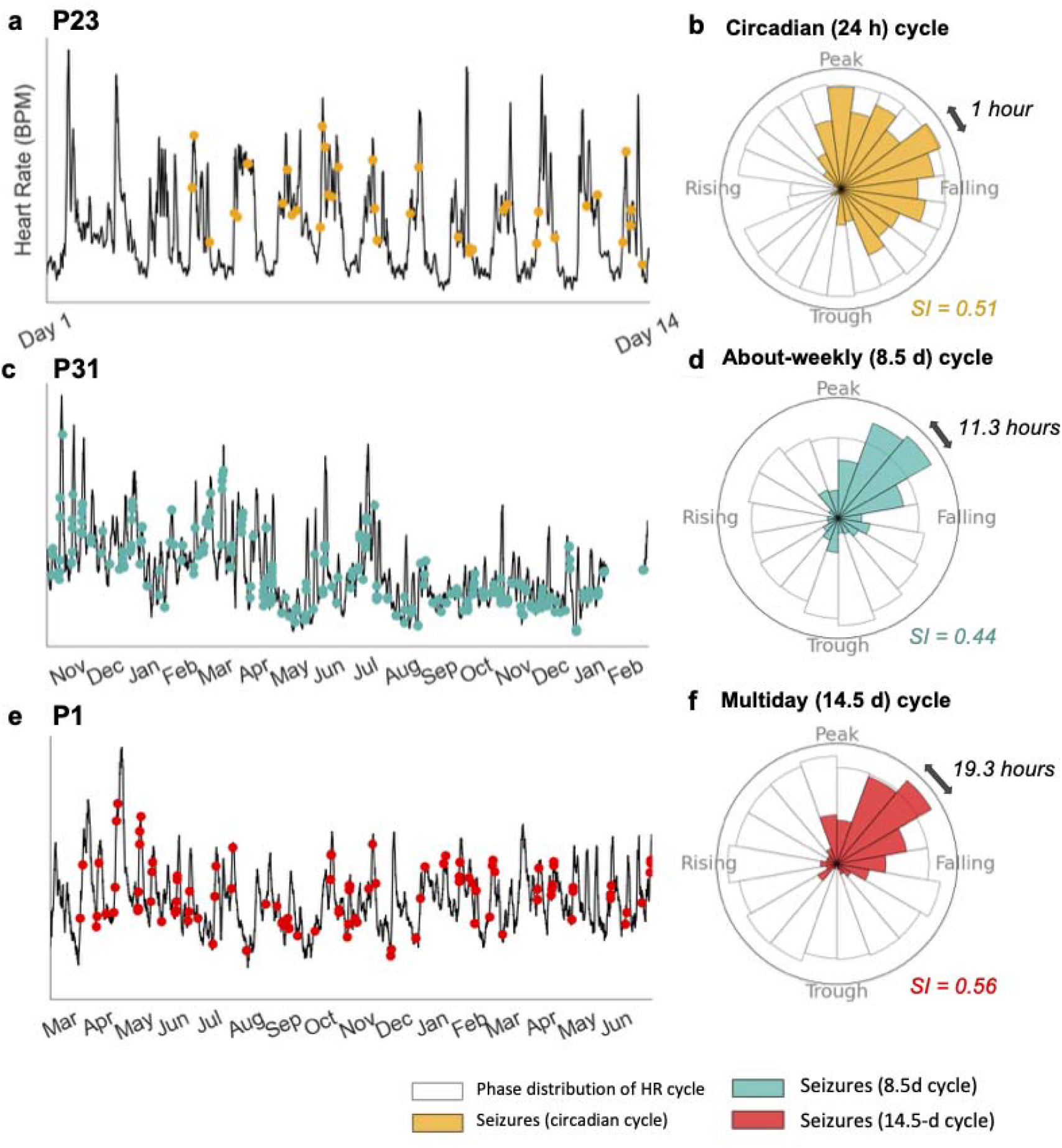
Examples of seizure occurrences locked to heart rate cycles for three participants. **(A, B)** P23 (416 seizures), **(C, D)** P31 (286 seizures), **(E, F)** P1 (105 seizures). **(A, C, E)** Heart rate (y-axis) and self-reported seizures (dots). A moving average (MA) filter was applied to heart rate (black line) to highlight cycles (**A:** 1-hour MA, **C, E:** 2-day MA). Panels (**B, D, F)** Corresponding circular histograms of the phase distributions of individuals’ heart rate cycles (white bins) showing the phase of seizure occurrences (shaded bins). Landmark phases are labelled as ‘peak’ (π/2), ‘trough’ (3π/2), ‘rising’ (2π) and ‘falling’ (π). Multiday circular histograms (Panels D, F) bins have the same phase width (2π/18) although these correspond to different durations (labelled by black arrows), depending on the period of the multiday cycle. The circadian histogram (Panel B) bins have widths of 1 hour (2π/24).

Figure 5 shows the mean resultant vectors for every heart rate cycle where significant seizure phase locking was observed (exact SI values, preferred phase/circular mean, and p-values are given in Supplementary Tables 4 and 5). For multiday cycles the phase of seizure occurrence appeared relatively diverse. For circadian cycles, seizures tended to occur on the falling phase (around afternoon); this distribution was suggestive of a reporting bias that could affect phase locking of self-reported events to circadian cycles (Supplementary Figure 6). However, it is important to note that a diurnal reporting bias would not affect phase-locking to multiday cycles.

**Figure 5.**
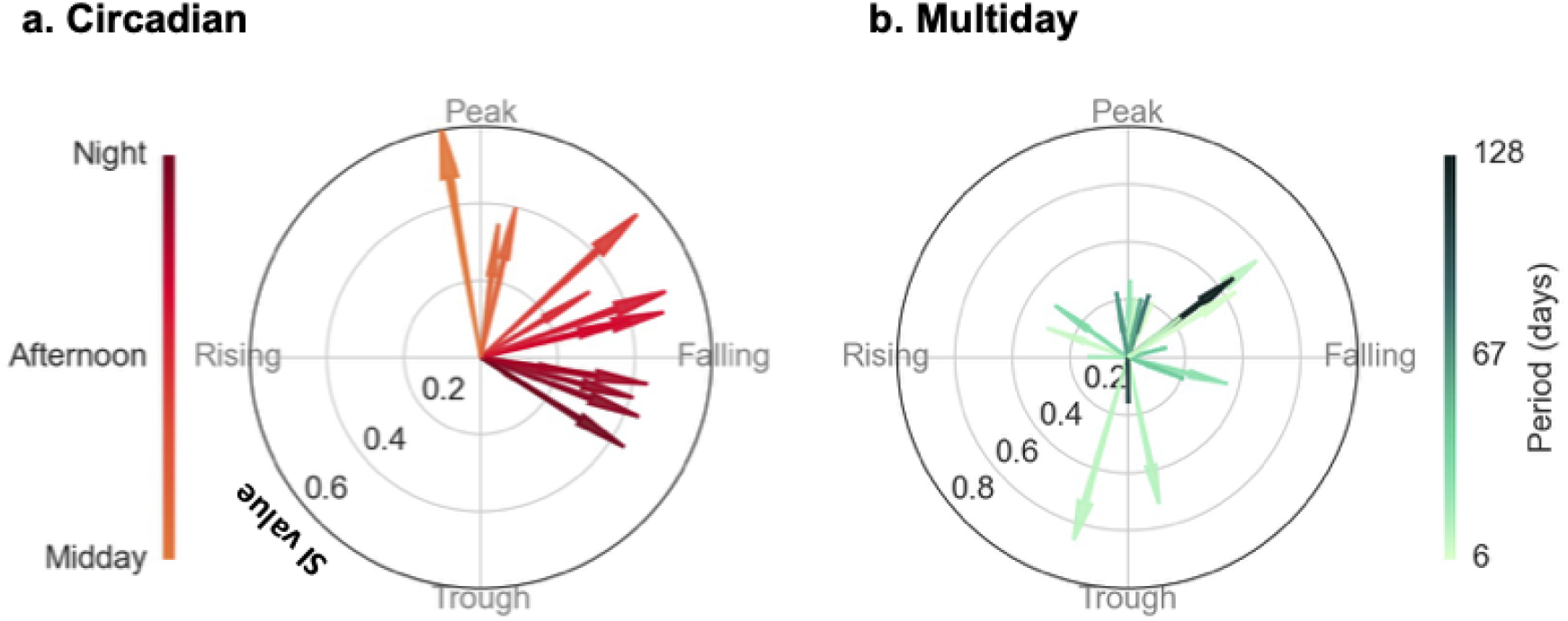
Phase locking of seizures to heart rate cycles. Both subplots show individual heart rate cycles (arrows) with significant phase locking of seizure occurrence. The lengths of the arrows indicate the strength of phase locking, or SI (radial axis, between 0 and 1), while the direction indicates the preferred phase of seizure occurrence (polar axis). **(A)** Circadian cycles, all periods were 24 hours. **(B)** Multiday cycles (including about-weekly and about-monthly), 6– to 128-day periods (colour bar).

## Discussion

Multiday heart rate cycles were found in most participants (N=29/31 people with epilepsy and N=12/15 healthy controls, Table 2), with striking examples of about-weekly and about-monthly rhythms, although not linked to the calendar week or month. Apart from a few limited studies^3^, such long-term, individual-specific rhythms in average heart rate have not previously been documented. Furthermore, for people with epilepsy, seizures preferentially occurred at limited phases of their multiday heart rate cycles in approximately half the individuals considered (10 out of 19), which is similar to the observed 60% prevalence of multiday epileptic rhythms in people with epilepsy.^11^ This phase relationship suggests heart rate cycles may be comodulated with already established multiday cycles of seizure likelihood. ^4–6^ Elucidating the relationship between periodic fluctuations in cortical excitability and heart rate has the potential to shed light on underlying mechanisms of multiday physiological rhythms with clinical applications for both neurological and cardiovascular diseases.

### Mechanisms of multiday cycles in the heart and brain

Endogenous multiday cycles of epileptic activity are well documented,^7^ with seminal studies in animal models^9,10^ and humans^5,8^ revealing free-running multiday cycles. Multiday heart rate cycles showed similar features to these cycles of epileptic brain activity, with cycles more common at about-weekly and about-monthly time scales (Figure 1), and a preference for seizures to occur on a particular phase of heart rate cycles (Figure 4, Table 2), which are also properties of epileptic rhythms.^5,6^ Nevertheless, the current study cannot determine whether multiday cycles in the brain and heart are causally linked. Ictal and peri-ictal increase in heart rate is common in epilepsy^21^, although the low resolution of self-reported seizure times prevented detection of ictal tachycardia in our study. In any case, acute, periictal heart rate changes would not account for the observed multiday cycles. Furthermore, the existence of similar rhythms in people without epilepsy (Figure 3) suggests that heart rate cycles were not primarily driven by the occurrence of seizures. Conversely, people without epilepsy likely do still experience fluctuations in brain excitability^22,23^. Circadian rhythms of heart rate are modulated via the autonomic nervous system and involve sympathetic-parasympathetic balance^24^, which is commonly disrupted in people with epilepsy^20^. Therefore, it is plausible that cortical and cardiac dynamics might also be linked over multiday timescales. Future investigations into a mechanistic relationship between multiday cycles of heart rate and epileptic activity are warranted.

In contrast to epileptology, the existence of multiday cycles of heart rate has not been widely described in the cardiology literature, although some earlier studies (limited to short-term recordings of less than one month, or case studies) have documented endogenous weekly rhythms and 7-day cycles in heart rate and blood pressure. ^17,18^ The current work also found multiday heart rate cycles commonly had weekly periodicities, in both men and women and people with and without epilepsy (Figures 1,3). Some about-weekly heart rate cycles were found with precisely 7-day periods (N=9/17 people with epilepsy and N=6/8 healthy controls), suggesting there was some entrainment by behavioural changes over the workweek. However, the occurrence of about-weekly cycles that were not linked to the workweek (i.e., between 5 – 9 days) indicates heart rate cycles also had an endogenous component.

The causal mechanisms of multiday epileptic rhythms or heart rate cycles are not understood, although several hormonal, metabolic and other environmental factors are implicated in endogenous multiday cycles in humans.^1–3,7^ Female sex hormones have been widely linked to monthly cycles of epileptic activity in women^25^ and the menstrual cycle may also lead to changes in heart rate and HRV.^26^ However, hormonal changes cannot explain the similar prevalence of monthly cycles of seizures in children and men,^4,5,27^ nor the present results demonstrating monthly cycles of heart rate in males (Figure 1c). Chronic stress can promote autonomic imbalance affecting both heart rate^28^ and epileptic brain activity^29^, and stress is also perceived as one of the leading factors triggering seizures^30^. However, stress is not typically considered to follow regular, multiday cycles. To elucidate the drivers of co-modulated cardiac and epileptic activity, future studies should target a range of candidate mechanisms for long-term monitoring.

### Clinical applications

Heart rate cycles can provide a biomarker for individual seizure likelihood, which may be used to guide epilepsy therapy or in seizure forecasting systems. Seizure forecasting is considered a key goal of epilepsy treatment, restoring a degree of control to people with refractory seizures; however, most forecasting algorithms have been deployed for intracranial EEG, which poses challenges for widespread availability of seizure warning systems.^31^ Importantly, forecasting algorithms based on multiday cycles have emerged as the most accurate, recently surpassing all previous approaches on a benchmark human dataset^6^ as well as showing excellent performance for larger cohorts,^32^ and using data from seizure diaries.^33^ If heart rate can be shown to reliably track multiday cycles of seizure likelihood, then similarly powerful seizure forecasts may eventually be derived from wearable devices, a concept that has already been demonstrated in retrospective studies^34,35^.

In addition to seizure forecasting, monitoring heart rate cycles may provide insight into the long-term role of heart rate and HRV in risk of sudden unexpected death in epilepsy (SUDEP). There are stereotypical cardiorespiratory changes prior to the occurrence of SUDEP^36^, with electrocardiographic abnormalities and impaired autonomic control associated with a higher risk of SUDEP.^37^ Long-term ECG monitoring may be predictive of SUDEP in people with epilepsy.^38,39^ However, despite circadian modulation of SUDEP risk - with substantially higher incidence at night^36^ - longer rhythms have not yet been investigated. The existence of co-modulated heart rate and seizure cycles makes heart rate cycles a feasible biomarker for the risk of both seizure occurrence and SUDEP.

Multiday heart rate cycles may have implications for cardiology. The relevance of circadian cycles to cardiovascular disease has long been recognised. Circadian patterns are observed across most arrhythmic events regardless of whether an underlying heart condition is present.^24^ In addition to circadian rhythms of cardiovascular disease, seven-day ^19^ and seasonal patterns ^40^ have been documented. Multiple studies show a Monday peak in cardiac mortality and hospital admissions for cardiovascular disease.^19^ Although it is hypothesised that these weekly patterns may be related to endogenous variation in cardiac output,^1^ weekday and seasonal trends are more likely to be driven by environmental factors.^19^ The current study found some precise 7-day heart rate cycles, although most multiday heart rate cycles were not locked to a 7-day week, or associated with a particular weekday, suggesting that incidence of cardiovascular disease should be explored in relation to underlying individual-specific cycle periods to investigate potential high-risk times for common dysfunctions. Just as circadian regulation leads to danger times for people with heart conditions, it is possible that risk factors for cardiac mortality also fluctuate over multiday timescales.

### Limitations and future work

The current study was based on self-reported seizure times, so noise could influence results. We had previously demonstrated that, for some people, multiday cycles established from seizure diaries align with cycles recorded from true electrographic seizures.^33^ Nevertheless, reporting bias could influence seizure timing, particularly with respect to circadian heart rate cycles, where reporting was likely to be affected by time of day (Supplementary Figure 6). On the other hand, weekly heart rate cycles were not always aligned to the precise 7-day workweek, so reporting bias linked to the day of the week would not drive synchronisation to these longer cycles (Supplementary Figure 7). This study did not monitor behavioural aspects such as exercise, medication adherence or dietary changes, which may drive multiday cycles.

Heart rate cycles were measured from a consumer, wearable device, with reduced accuracy and temporal resolution compared to ECG. These limitations restricted the assessment of cardiac signals to look at average heart rate (within a 5-minute window), rather than other features, such as HRV, which may be critical to understanding brain-heart interactions in epilepsy.^20^ Wearable heart rate sensors are also subject to artefacts, although measurement noise was likely to be at a higher frequency than the multiday time scale focused on in the current work. It is worth noting that the heart rate measured via Fitbit photoplethysmography has been validated against ECG, showing no significant difference in resting heart rate during sleep ^42^, although systematic errors emerge during high intensity exercise.^43^

Future work will extend these results to validate the existence of multiday heart rate cycles and their relationship to electrographic seizures using chronic sub-scalp EEG and additional wearable sensors. For now, it is our hope that the prospect of multiday cycles governing diverse physiological systems leads to new breakthroughs in understanding biological rhythms and treatment of disease.

## Methods

### Study design

Tracking Seizure Cycles is an ongoing observational cohort study examining seizure and other biological cycles and their interactions using long-term non-invasive monitoring. The study was approved by the St Vincent’s Hospital Human Research Ethics Committee (HREC 009.19).

### Participants

Adults (18 years and over) with a confirmed epilepsy diagnosis and healthy controls were recruited between August 2019 and March 2021. Participants with epilepsy had uncontrolled or partially controlled seizures and were recruited through neurologist referral. Healthy controls were recruited from the general population. All participants provided written informed consent.

### Data collection

Continuous data were collected via mobile and wearable devices. All participants wore a smartwatch (Fitbit) and participants with epilepsy manually reported seizure times in a freely available mobile diary app (Seer App). Participants with epilepsy were instructed to report all their clinically apparent events, including generalised and focal seizures (both aware and unaware). The smartwatch continuously measured participants’ heart rates (via photoplethysmography) at 5s resolution.

### Statistical analyses

All analyses were executed in Python (version 3.7.9).

#### Heart rate cycle analysis

To evaluate the strength of heart rate cycles, participants were required to have at least four months of recordings with over 80% adherence (i.e., less than 20% missing data), with adherence defined as at least one heart rate recording every hour.

Cycles were measured at different periods using a wavelet transform approach that was previously proposed for the detection of multiday cycles of epileptic activity.^5^ The continuous heart rate signal was first down-sampled to one timestamp every five minutes. Linear interpolation was performed for up to 1 hour either side of a missing segment, to account for time to charge the wearable device. Longer recording gaps were interpolated with a straight line at the average value of all the data (see Supplementary Appendix 1). Missing segments that were interpolated with a straight line ranged from 0.1 - 1354 hours (M = 17.9, SD = 68.3 hours). Simulation showed that most cycles could be detected after interpolation, even with long missing segments, or multiple missing segments (Supplementary Table 3 and Supplementary Figure 2).

Prior to wavelet analysis the heart rate signal was z-standardised (by subtracting the mean and dividing by the standard deviation). Candidate cycle periods were then tested using a continuous Morlet wavelet transform, where significant peaks in the global wavelet power (using the time-averaged significance test described in ^44^) were determined to be significant heart rate cycle periods (Supplementary Appendix 2). We considered the cone of influence of the wavelet – areas of the wavelet that are potentially affected by edge-effect artefacts – by restricting cycle periods from 2.4 hours to a maximum period of one-quarter of the recording length (i.e., a minimum of four cycles had to be observed). A Morlet wavelet was chosen to be consistent with other multiday cycles work in epilepsy^5^. We also found the wavelet approach to be less vulnerable to noise than Fourier transformations, although both methods did pick up multiple closely spaced peaks (Supplementary Figure 4), likely because physiological data does not display perfect periodicity. To eliminate some of these additional peaks we used a peak significance threshold of 99%.

To extract heart rate cycles, the standardised heart rate signal was bandpass filtered into distinct component frequencies matching the significant cycle frequencies (identified from wavelet decomposition). The bandpass filter applied at each significant cycle was a second-order zero-phase Butterworth bandpass filter with cut-off frequencies at ±33.3% of the cycle frequency. For instance, someone with significant cycles (wavelet spectrum peaks) at 24 hours, 9 days and 30 days would have three bandpass filters applied with cut-off frequencies of 16-32 hours, 6-12 days and 20-40 days, respectively. These cut-off frequencies were chosen to account for phase shifts in the cycle over the recording time and is consistent with previous work.^5^ To account for bandpass overlap in significant cycle frequencies, we introduced a sparsity criterion, whereby only the strongest peak within any cycle’s bandpass filter pass band was considered.

#### Relationship between seizures and heart rate cycles

To evaluate potential co-modulation between seizure occurrence and heart rate cycles, participants were required to have at least 20 reported seizures. Co-modulation was measured from the degree of ‘phase-locking’ of seizure times with respect to underlying heart rate cycles, i.e., how tightly seizure times were linked to a particular point (phase) of the cycle. Phase-locking was quantified by the magnitude of the mean resultant vector computed from the phase of all seizures. The magnitude of the mean resultant vector (henceforth called the synchronisation index, SI) ranges from 0 to 1, where 0 indicates a uniform distribution (i.e., no relationship) and 1 indicates perfect alignment with respect to an underlying cycle. The angle or direction of the mean resultant vector indicates the preferred phase of seizure occurrence; for instance, seizures could be more likely near the peak or trough of average heart rate cycles.

The continuous phase of heart rate cycles was estimated using the Hilbert transform. The times of seizure occurrence were mapped to the estimated phase of heart rate cycles. Seizure phases were then binned into 24 (circadian cycles) or 18 (all other periods) equal sized bins (ranging from 0 to 2π) to produce a phase distribution. The SI values were computed from these phase distributions. An omnibus (Hodges-Ajne) test^45^ was used to determine whether seizures were significantly phase-locked to the heart rate cycle by testing the null hypothesis that the phase distribution was uniform. A Bonferroni correction was used to reduce the experiment-wise error, by accounting for comparisons across multiple heart rate cycles (see Supplementary Appendix 3).

## Data Availability

Data will be made available upon completion of the study (2022). Data used in this manuscript can be made available upon reasonable request to the corresponding author.

## Data availability

Excluding participants who did not consent to share their data publicly, all deidentified data will be made available on Figshare following publication.

