## Supplementary for "Multiday cycles of heart rate are associated with seizure likelihood"

### Supplement

**Supplementary Table 1. AEDs of eligible participants in Tracking Seizure Cycles cohort**

| Participant | Anti-Epileptic Medication |
|---|---|
| P1 | Levetiracetam, Lamotrigine |
| P2 | Levetiracetam, Topiramate, Lamotrigine |
| P3 | Sodium Valproate |
| P4 | Levetiracetam, Lacosamide |
| P5 | None |
| P6 | Lamotrigine, Clonazepam, Oxcarbazepine, Zonisamide |
| P7 | Clobazam, Lacosamide, Levetiracetam, Oxcarbazepine |
| P8 | Brivaracetam, Phenobarbital, Clonazepam, Carbamazepine |
| P9 | Lacosamide, Zonisamide, Clobazam |
| P10 | Sodium Valproate, Lamotrigine |
| P11 | Sodium Valproate, Lamotrigine, Perampanel |
| P12 | Levetiracetam, Topiramate |
| P13 | Zonisamide, Lacosamide |
| P14 | Brivaracetam, Levetiracetam |
| P15 | Levetiracetam, Topiramate, Lacosamide |
| P16 | Sodium Valproate, Pregabalin, Lacosamide |
| P17 | Topiramate, Lamotrigine, Carbamazepine |
| P18 | Carbamazepine |
| P19 | Sodium Valproate, Pregabalin, Lacosamide |
| P20 | Levetiracetam, Topiramate, Lamotrigine |
| P21 | Brivaracetam, Lacosamide, Perampanel, Lamotrigine |
| P22 | Sodium Valproate |
| P23 | Levetiracetam, Topiramate, Lacosamide |
| P24 | Zonisamide, Lamotrigine, Clonazapam, Medicinal Cannabis |
| P25 | Sodium Valproate, Lamotrigine, Perampanel |
| P26 | Lacosamide, Brivaracetam, Zonisamide, Losartan |
| P27 | Oxcarbazepine |
| P28 | Carbamazepine, Clobazam |

**Supplementary Table 2. Control participants in Tracking Seizure Cycles cohort**

| Participant | Gender | Data duration (months) | Adherence (%) | Average Heart Rate (BPM) |
|---|---|---|---|---|
| C1 | F | 6.7 | 93 | 70 |
| C2 | M | 5.5 | 97 | 70 |
| C3 | F | 16.6 | 80 | 71 |
| C4 | F | 6.2 | 83 | 75 |
| C5 | F | 6.5 | 97 | 77 |
| C6 | M | 6.8 | 97 | 67 |
| C7 | M | 5.5 | 97 | 67 |
| C8 | F | 17 | 91 | 72 |
| C9 | M | 26.2 | 97 | 69 |
| C10 | F | 6.3 | 92 | 78 |
| C11 | M | 6 | 99 | 61 |
| C12 | M | 6.2 | 93 | 75 |
| C13 | M | 5.9 | 96 | 69 |
| C14 | M | 6.8 | 80 | 64 |

#### Appendix 1: Heart rate data interpolation

To select the best interpolation method for missing data segments, we chose a participant (P14) with no missing data segments, found their true heart rate cycles, and then arbitrarily removed up to 20% of the raw data (Supplementary Figure 1a). Following data removal, three different interpolation methods were applied to missing segments (Supplementary

Figure 1b), cycles were detected using the Morlet wavelet and resolved cycles across interpolation methods were compared to the true cycles.

Three interpolation methods were applied to the missing data segments: ‘average time of day’ method, ‘straight line’ method and ‘copy’ method. The ‘average time of day’ method interpolated each data point with the time-matched average heart rate (e.g., a missing data point at 9AM was replaced with the average heart rate at 9AM across the whole dataset). The ‘straight line’ method interpolated each data point with the average heart rate found across the whole dataset. The ‘copy’ method interpolated each data point with the time-matched most recent heart rate values (e.g., a missing data segment from 9AM-3PM on January 1, 2020 was replaced with the data from 9AM-3PM on December 31, 2019).

To compare the interpolation methods, the Morlet wavelet was used to determine significant cycles in the data. Cycles were compared among interpolation methods to the true cycles (original data peaks detected in the Morlet wavelet spectrum). After 1000 runs of arbitrary data removal, 92.1%, 93.5% and 95.9% of true cycles were resolved and 0.40, 0.49 and 0.86 extra peaks were detected using the ‘average time of day’, ‘straight line’ and ‘copy’ methods, respectively. The ‘straight line’ method was chosen for its simplicity and overall performance while minimising erroneous cycle detections.

To further investigate the effect of the ‘straight line’ interpolation on cycle detection, we performed another simulation on three datasets from two other participants (P4 and P26), where a four-month period existed with no missing data. To simulate the worst-case scenario, we systematically removed between 1 and 30 segments (increasing by one segment each simulation), where deleted segment lengths were arbitrarily chosen such that the total removed time was always 20% of the total recording time. The results of simulation are shown in Supplementary Table 3. On average over 75% of actual peaks (cycles) were detected despite interpolation. Two example simulations are shown in Supplementary Figure 2, where one large segment (20% of the data length) and 15 shorter segments (arbitrary lengths totalling 20% of the data length) were removed. Both simulations retrieved cycles (Supplementary Figs 2d and 2f) that were similar to the true wavelet (Supplementary Figure 2b). The simulation of one removed segment detected cycles that were similar to the true cycles, despite a large missing chunk. However, the simulation of 15 removed chucks missed a true cycle at 72h, which was much weaker than the other peaks in the original wavelet. Overall, stronger peaks in the initial wavelet were typically detected in all simulations (regardless of length and number of missing segments), and weaker peaks were more vulnerable to going undetected.

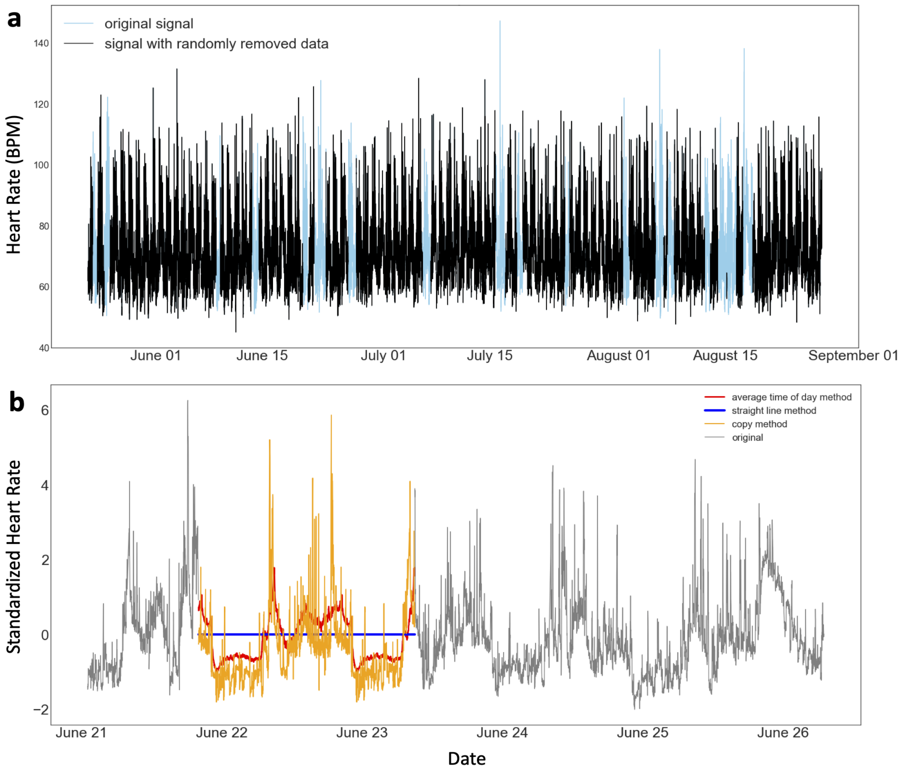


**Supplementary Figure 1. Interpolated data simulation results for participant P14. a:** Example simulation where up to 20% of the data was arbitrarily removed. **b:** Example missing segments with three interpolation methods: average time of day method (red), straight line method (blue) and copy method (orange).

**Supplementary Table 3. Results of interpolated data simulation experiment on three segments without missing data.** Two participants (P4 and P24) were used who had data segments of at least four months with no missing segments (note that P4 had two, non-overlapping four-month segments with no missing data)

| | **Segment duration (days)** | **Total time removed in simulation (hours)** | **Average true peak resolution (SD)** |
|---|---|---|---|
| **P4** | 208 | 998 | 84% (12%) |
| **P4** | 146 | 698 | 79% (19%) |
| **P24** | 273 | 1308 | 75% (17%) |

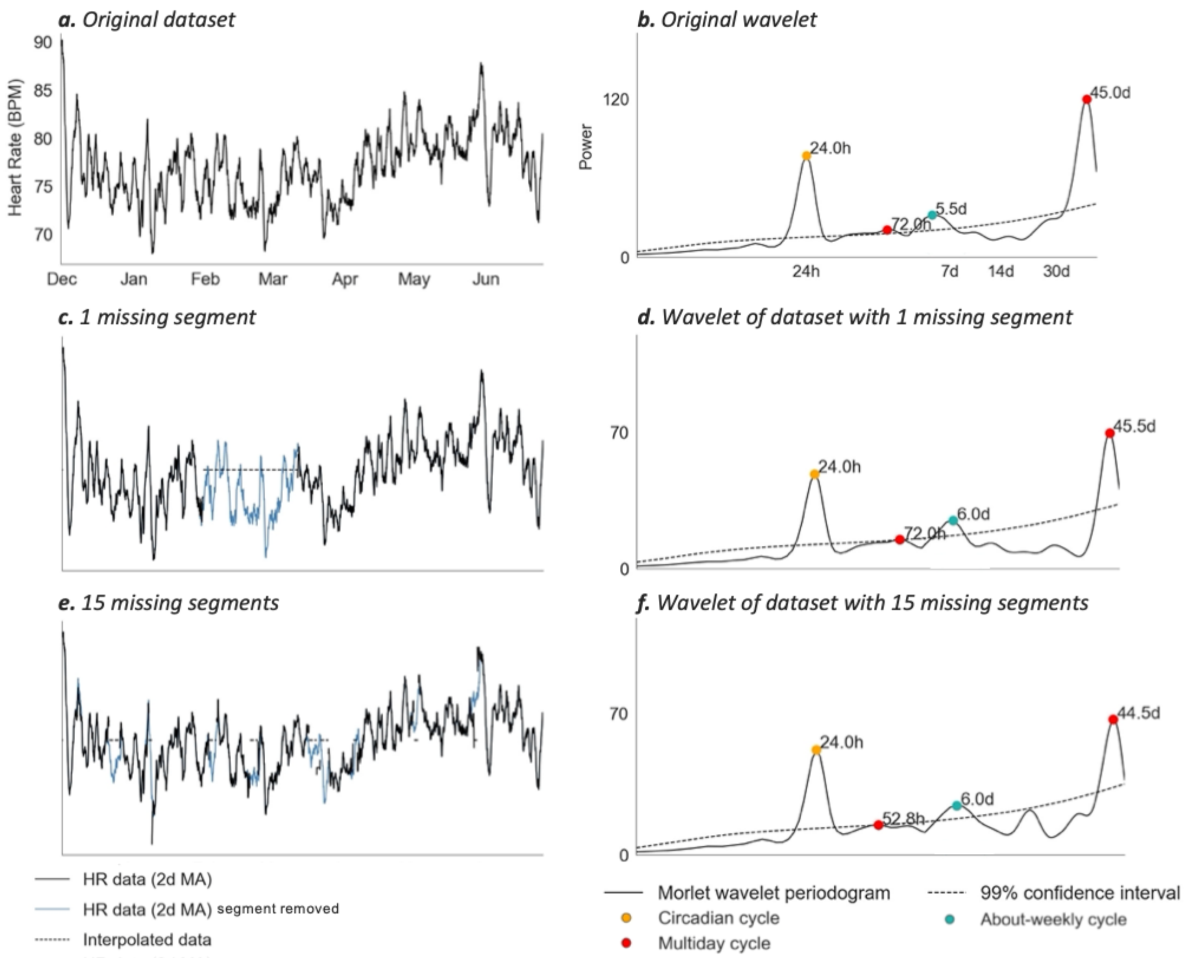


**Supplementary Figure 2. Changes in cycles detected in P4's wavelet spectrum after data removed and interpolated. a:** Heart rate (HR) data segment (208 days) with no missing data **b:** Wavelet spectrum of original dataset. **c,d:** HR data after segments (**c:**1 segment of 998 hours, **d:**15 segments totalling 998 hours) were arbitrarily removed. Removed segments are shown in blue and interpolation values are given by the dotted line. **e,f:** Wavelets after data was removed and interpolated with the average straight line. Note the similarly in cycles (wavelet peaks) retrieved after large segments of data are interpolated.

**Appendix 2: Detection of multiday heart rate cycles**

Prior to wavelet analysis the heart rate signal (post-interpolation) was z-standardised (by subtracting the mean and dividing by the standard deviation). A continuous Morlet wavelet transform with increased spacing was used on standardised data segments to compute the power at different scales (cycle periods).[1] The scales were every 1.2 hours between 2.4 and 31.2 hours, every 2.4 hours between 33.6 and 48 hours, every 4.8 hours between 52.8 and 4 days and every 12 hours between 5 days and up to a maximum period of one quarter of the recording duration. The data were then represented as a global wavelet spectrum of power (the product of the average of the square of absolute value of complex wavelet coefficients and the variance of the time series) for each scale (cycle). Peaks in the wavelet spectrum were found by comparing neighbouring values. Peaks above the global significance (99% confidence) level were determined to be significant heart rate cycle periods using a time-averaged significance test (Torrence and Compo, 1998).

Supplementary Figure 3 shows heart rate recordings and significant detected heart rate cycles (wavelet spectrum) for all eligible participants. Supplementary Figure 4 shows the wavelet spectrums of each participant overlaid with the Fast Fourier Transform (FFT) spectrum for each participant. Supplementary Figure 5 shows an example of heart rate cycles for two people in the control cohort.

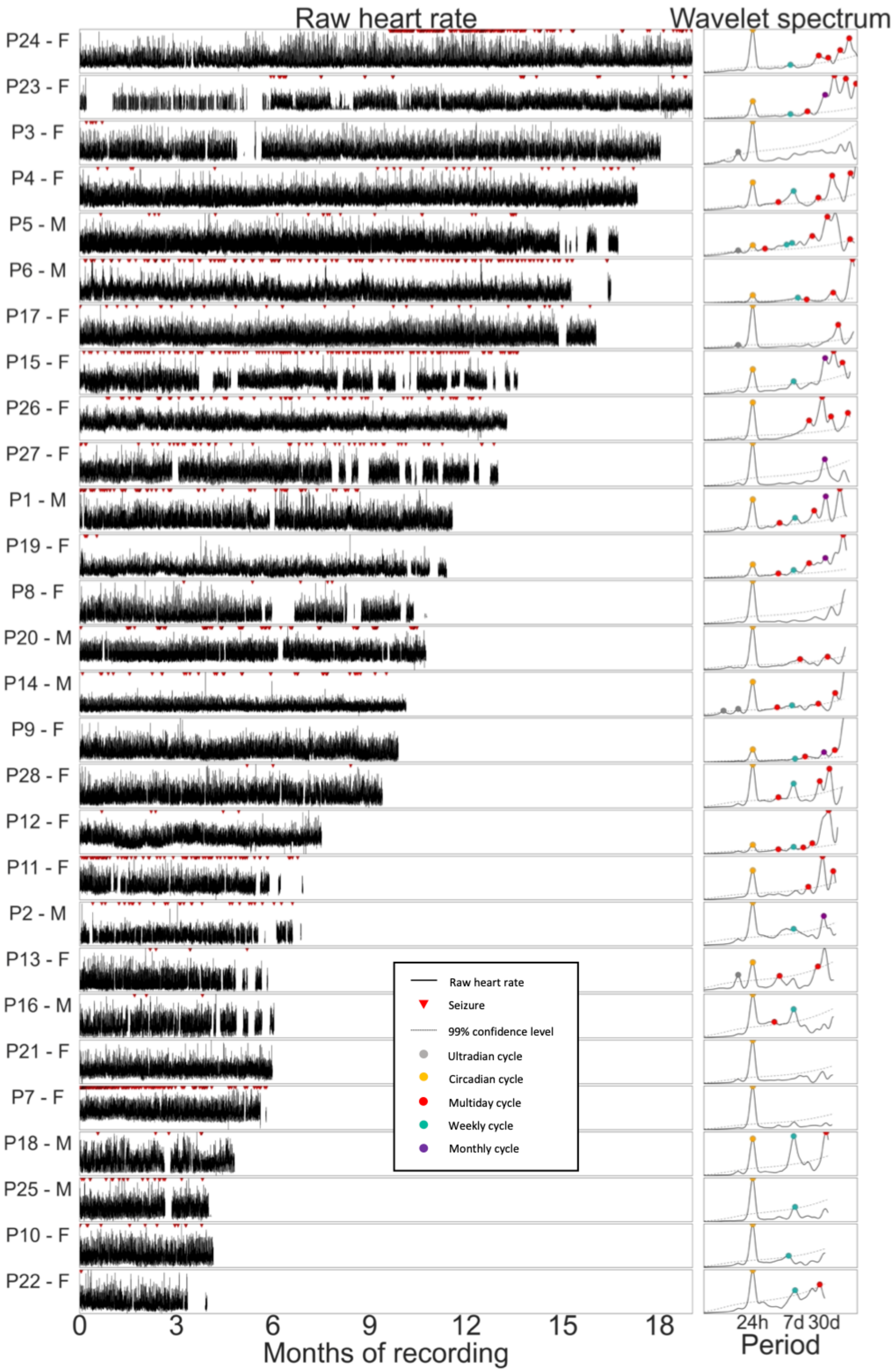


**Supplementary Figure 3. Individual raw heart rate recordings and wavelet spectra for 28 participants with epilepsy.** Raw heart rate data (black) are shown on the left, with seizures shown as red triangles at the top. Wavelet spectra are shown on the right with significant cycles labelled: ultradian (grey), circadian (orange), multiday (red), weekly (turquoise) and monthly (purple). Global confidence levels are represented on the wavelet spectra by dashed black lines.

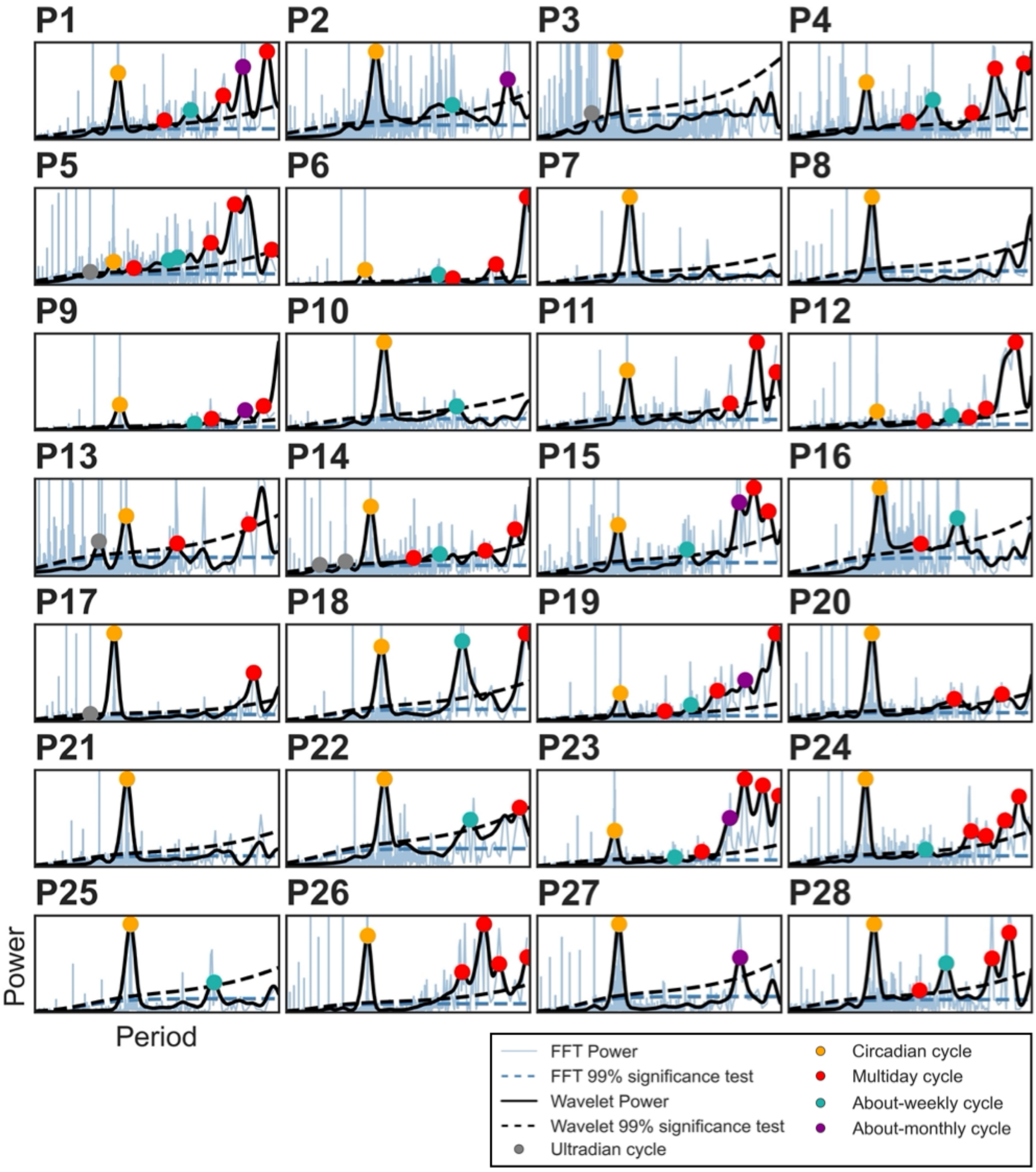


**Supplementary Figure 4. Individual wavelet and Fast Fourier Transform (FFT) spectra for 28 eligible participants with epilepsy.** Wavelet spectra (black solid lines) are shown with significant cycles labelled (coloured dots): ultradian (grey), circadian (orange), multiday (red), weekly (turquoise) and monthly (purple). FFT spectra are represented by the solid blue lines. Peaks in wavelet spectra correspond to peaks in FFT spectra, but wavelet spectra typically have fewer significant peaks. Global significance levels (99% confidence) are shown for both the wavelet (dashed black lines) and FFT spectra (dashed blue lines).

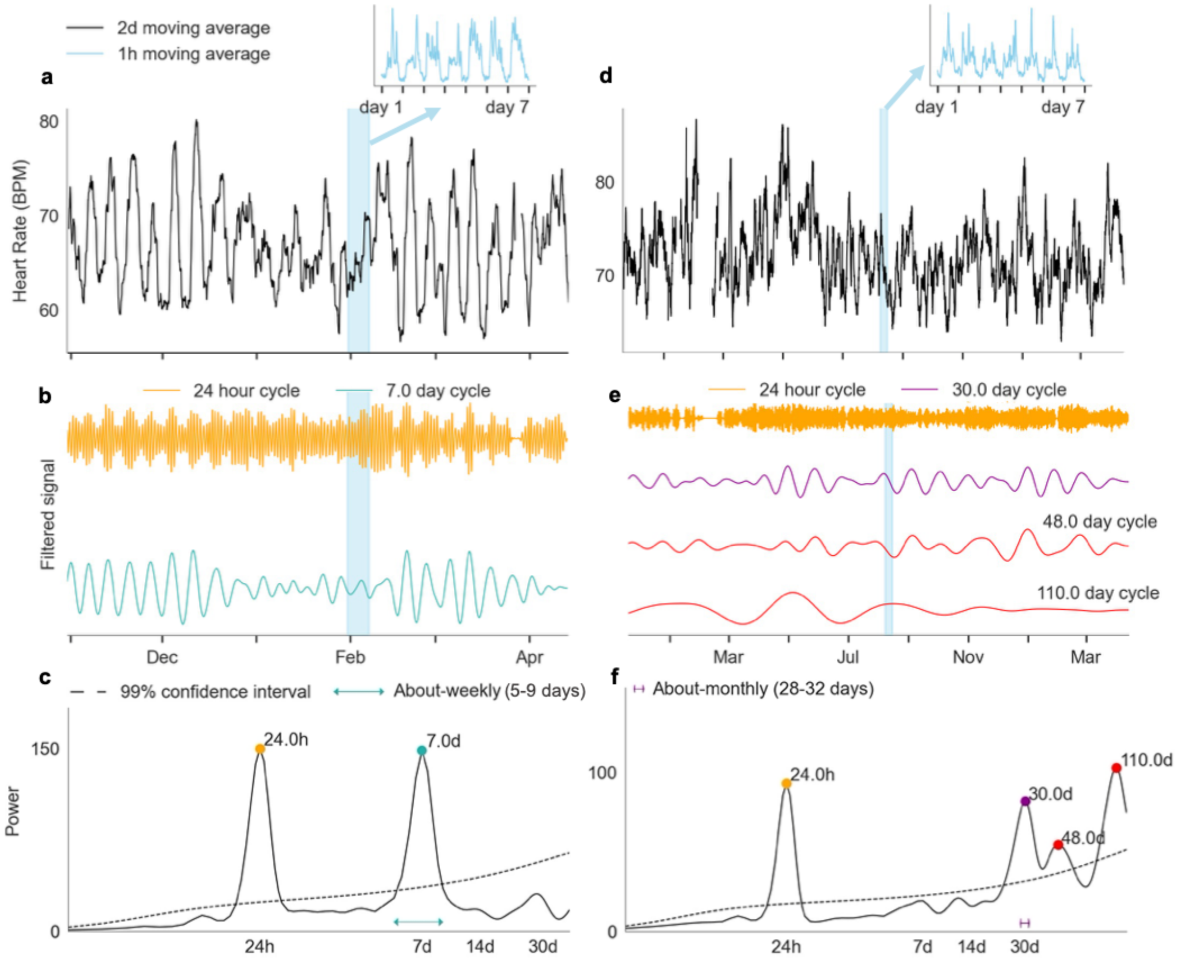


**Supplementary Figure 5. Examples of multiday heart rate cycles in people without epilepsy.** Data are shown for two different participants without epilepsy, C7 (a-c) and C8 (d-f). **a,d:** Heart rate (y-axis) smoothed with a 2-day moving average filter shows multiday cycles. Insets (blue) show circadian rhythms of heart rate. **b,e:** A graphical representation of the bandpass filtered heart rate signals for different cycles (corresponding respectively to spectrum peaks in panels c,f). Note that the signal amplitudes for different cycles (coloured traces) have been normalised to have the same range. **c,f:** Wavelet power spectra for different scales (x-axis). Significant cycle periods (peaks) are labelled with coloured dots.

#### Appendix 3: Phase locking of seizures to heart rate cycles

An omnibus (Hodges-Ajne) test was used to determine whether seizures were significantly phase-locked to the heart rate cycle by testing the null hypothesis that the phase distribution was uniform. The Bonferroni correction was used to account for comparisons across multiple heart rate cycles. Supplementary Tables 4 and 5 show Bonferroni corrected and observed p-values for individuals' cycles (multiday and circadian, respectively), as well as the observed SI, circular mean (i.e., the angle of the mean resultant vector, indicating the direction of phase locking).

**Supplementary Table 4. Multiday heart rate cycles where seizures are significantly phase locked.** Each cycle is shown with the p-value required for significance (after Bonferroni adjustment for multiple comparisons test on p = 0.05), the p-value of the seizure distribution (always less than the p-value required for significance), the SI value and the circular mean (between 0 and 2π).

| Participant | Cycle (hours) | Peak type | P-value required | P-value of seizure distribution | SI value of seizure distribution | Circular mean (radians) |
|---|---|---|---|---|---|---|
| **1** | 180 | weekly | 0.0083 | 0.0039 | 0.21 | 2 |
| **5** | 408 | multiday | 0.0063 | 0.00066 | 0.54 | 4.3 |
| | 1092 | multiday | 0.01 | 0.002 | 0.14 | 2.9 |
| | 204 | weekly | 0.01 | 4.10E-17 | 0.44 | 2.6 |
| | 2712 | multiday | 0.01 | 3.30E-06 | 0.16 | 4.7 |
| **6** | 312 | multiday | 0.01 | 6.00E-06 | 0.23 | 2.5 |
| **14** | 540 | multiday | 0.0071 | 0.0019 | 0.36 | 4.4 |
| **15** | 1692 | multiday | 0.01 | 0.0083 | 0.23 | 1.4 |
| | 228 | multiday | 0.017 | 6.70E-10 | 0.4 | 0.44 |
| **20** | 840 | multiday | 0.017 | 6.70E-12 | 0.42 | 0.53 |
| | 1152 | multiday | 0.0071 | 3.00E-05 | 0.25 | 3.7 |
| | 324 | multiday | 0.0071 | 2.80E-07 | 0.34 | 1.8 |
| | 744 | monthly | 0.0071 | 2.80E-07 | 0.46 | 3.4 |
| | 144 | weekly | 0.0071 | 1.00E-05 | 0.34 | 2 |
| **23** | 3228 | multiday | 0.0071 | 2.00E-08 | 0.43 | 2.5 |
| | 552 | multiday | 0.0083 | 0.0001 | 0.17 | 6 |
| | 144 | weekly | 0.0083 | 0.0019 | 0.07 | 1.5 |
| | 1524 | multiday | 0.0083 | 0.0041 | 0.12 | 0.71 |
| **24** | 2328 | multiday | 0.0083 | 0.0019 | 0.13 | 5.7 |
| **26** | 348 | multiday | 0.01 | 2.70E-07 | 0.49 | 2.5 |

**Supplementary Table 5. Circadian and ultradian heart rate cycles where seizures are significantly phase locked.** Each cycle is shown with the p-value required for significance (after Bonferroni adjustment for multiple comparisons test on p = 0.05), the p-value of the seizure distribution (always less than the p-value required for significance), the SI value and the circular mean (between 0 and 2π).

| Participant | Cycle (hours) | Peak type | P-value required | P-value of seizure distribution | SI value of seizure distribution | Circular mean (radians) |
|---|---|---|---|---|---|---|
| **1** | 24 | circadian | 0.0083 | 6.00E-17 | 0.56 | 2.4 |
| **2** | 24 | circadian | 0.017 | 0.0084 | 0.4 | 2.9 |
| **6** | 24 | circadian | 0.01 | 6.60E-15 | 0.4 | 1.8 |
| **7** | 24 | circadian | 0.05 | 1.10E-08 | 0.31 | 3.3 |
| **11** | 24 | circadian | 0.013 | 9.30E-18 | 0.44 | 3.5 |
| **14** | 6 | ultradian | 0.0071 | 0.00014 | 0.44 | 2.4 |

| | | | | | | |
|---|---|---|---|---|---|---|
| | 12 | ultradian | 0.0071 | 0.00055 | 0.41 | 3 |
| **15** | 24 | circadian | 0.01 | 4.60E-12 | 0.44 | 3.7 |
| **17** | 24 | circadian | 0.017 | 0.00031 | 0.46 | 2.8 |
| **20** | 24 | circadian | 0.017 | 0.00083 | 0.18 | 3.5 |
| **23** | 24 | circadian | 0.0071 | 9.60E-07 | 0.34 | 2.3 |
| **24** | 24 | circadian | 0.0083 | 2.80E-43 | 0.52 | 2.9 |
| **26** | 24 | circadian | 0.01 | 2.70E-05 | 0.37 | 3.4 |
| **27** | 24 | circadian | 0.025 | 0.0016 | 0.38 | 1.8 |

**Appendix 4: Phase locking to time of day and day of week**

In addition to the effect size of phase-locking we considered potential biases by entrainment of seizures to time-of-day and day-of-week. The circadian modulation of seizure occurrence could not easily be differentiated between circadian cycles of heart rate versus clock time. Supplementary Figure 6 shows phase locking of self-reported seizures with respect to time of day in the cohort of 16 people with more than 20 seizures. 71% of seizures were reported during 8AM and 8PM.

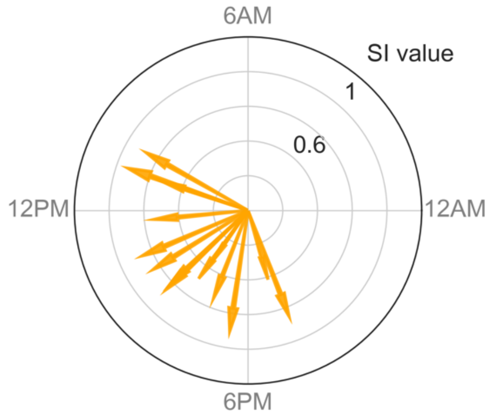


**Supplementary Figure 6. Phase locking of self-reported seizures to time of day.** Each arrow represents an individual (cohort of people with more than 20 seizures, N=16) with significant phase locking of their self-reported seizures to a particular time of day. The length of the arrows represents the strength of the phase locking (synchronization index, SI, value between 0 and 1) and the directions of the arrows indicate the peak times of seizure occurrence.

The tendency of seizures to occur on a particular day of the weekly heart rate cycle was also studied. Supplementary Figure 7 shows the empirical probabilities of seizures occurring on a particular day of the week for people with precise 7-day heart rate cycles (N=8). There was no evident trend for a preferred day/s on which more seizures were reported.

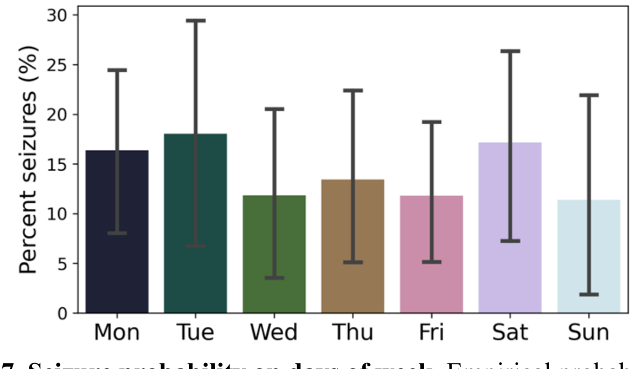


**Supplementary Figure 7. Seizure probability on days of week.** Empirical probabilities from seizure diaries are given for the cohort of people with precisely 7-day heart rate cycles (N = 8). Empirical probabilities shown are the mean and standard error of normalized individual seizure distributions across the population. Normalizing the individual distributions ensured people with more seizures did not bias the results.
